# Oxygen saturation and acute mountain sickness during repeated altitude exposures simulating high-altitude working schedules

**DOI:** 10.1101/2024.12.20.24319405

**Authors:** N. Waldner, S. E. Hartmann, L. Muralt, M. Lichtblau, P. R. Bader, J. M. Rawling, I. Lopez, S. Ulrich, M. J. Poulin, K.E. Bloch, M. Furian

## Abstract

**Objective:** To quantify the effect of two consecutive prolonged, intermittent exposures to high and very high altitudes on oxygen saturation (SpO_2_) and acute mountain sickness (AMS).

**Methods:** Healthy lowlanders (N=21), aged 18-30y, underwent two 7-day sojourns at the ALMA observatory, Chile (6hrs/day at 5050m, 18hrs/day at 2900m), separated by 1-week at 520m. SpO_2_ (pulse oximetry) and AMS severity (AMSc, Environmental Symptom Questionnaire cerebral score) diagnosing AMS (AMSc≥0.7) were assessed daily at both altitudes. www.ClinicalTrials.gov:NCT02730143.

**Results:** SpO_2_ at 2900m and 5050m on arrival day was mean±SD 93.6±0.5% and 79.9±1.0% (P<0.05 between altitudes), whereas the AMSc scores were 0.43±0.08 and 0.97±0.11 (P<0.05 between altitudes), respectively. At 2900m during a 7-day intermittent hypoxic exposure, SpO_2_ increased by a mean(95%CI) 0.3%/day(0.1;0.4) and by 0.9%/day(0.4;1.3) at 5050m. Similarly, AMSc decreased by 0.05points/day(0.01;0.08) at 2900m and by 0.16points/day(0.11;0.21) at 5050m. During the second sojourn (vs. 1^st^ sojourn), day1, SpO_2_ at 2900m was unchanged but higher at 5050m by 2.9%(0.6;5.3). AMSc was lower at 2900m and 5050m by 0.37(0.16;0.59) and 0.37(0.11;0.63) (P<0.05 both comparisons vs 1^st^ sojourn), respectively. Acclimatization with the 2^nd^ sojourn resulted in an increase in SpO_2_ at 2900m by 0.3%/day(0.1;0.4) and at 5050m: 0.5%/day(0.1;0.8). AMSc remained unchanged with acclimatization at 2900m but decreased at 5050m by 0.08points/day(0.04;0.11).

**Conclusions:** In healthy lowlanders, a 7-day intermittent hypobaric hypoxic exposure of varying severity at high and very high altitude improved SpO_2_ and AMS severity at 2900m, with larger improvements at 5050m. During a second identical sojourn, initial AMS severity was reduced despite comparable SpO_2_ at 2900m compared to the 1^st^ sojourn. No further acclimatization effects were observed in SpO_2_ but in AMS symptoms at 2900m. In contrast, re-exposure to 5050m showed higher initial SpO_2_ and lower AMSc values with further improvement with intermittent re-exposures. These findings highlight altitude-dependent acclimatization patterns and confirm the effectiveness of pre-conditioning to prevent AMS when planning sojourns to high altitudes.

## 1. Introduction

Various infrastructures are located at high altitude especially for tourism, mining industry, military operations, and astronomical observation centers. For example, workers of the Atacama Large Millimeter/submillimeter Array (ALMA) on the Chajnantor Plateau in Chile spend their nights at 2900 m (high altitude) and daytime work at 5050 m (very high altitude) for 7 days, followed by 7 days of recovery close to sea-level (Savioli et al., 2022). Due to their typical working schedule, rapid ascents with intermittent hypobaric hypoxic exposure to different severity of hypoxia are necessary, which makes acclimatization difficult (Furian et al., 2022b; Graf et al., 2022; Pun, Hartmann, et al., 2018).Acute exposure to high altitude is well-known to cause hypoxemia with potential development of acute mountain sickness (AMS) symptoms such as headache, dizziness, weakness, and nausea (Luks et al., 2019). Previous studies show that subjects who are already acclimatized to higher altitudes have a faster rise in arterial oxygen saturation (SpO_2_) after ascent to altitude compared to none-acclimatized sea-level residents when ascending to high altitude (Reeves et al., 1993). Acclimatization in form of intermittent altitude exposure improves SpO_2_ and reduces incidence and severity of AMS and therefore may be used as an alternative to prolonged altitude residence (Beidleman et al., 2004; Subudhi, Bourdillon, et al., 2014; Subudhi, Fan, et al., 2014).

The previously observed benefits of acclimatization through intermittent altitude exposure on AMS and SpO_2_ have not been thoroughly examined to determine whether they depend on the target altitude. Additionally, it remains unclear if these effects are also present in the intermittent altitude exposure sequences commonly experienced by high-altitude workers.

Therefore, this study aimed to evaluate the physiological effects of acute, prolonged intermittent hypobaric hypoxic exposure, and re-exposure on SpO_2_, AMS symptoms and vital parameters in healthy, not acclimatized participants. This protocol mimicked typical working schedule rotations of workers at ALMA and many other high-altitude work sites worldwide, i.e., 7 work days at high and very high altitude alternating with 7 leisure days near sea-level.

This study has three hypotheses. First, during acute high-altitude exposure, SpO_2_ will be reduced, and AMS symptoms will be pronounced compared to 520 m. Second, intermittent exposure to altitude during a 7-day stay that consists of sleeping and working altitude of 2900 and 5050 m, respectively, will improve SpO_2_ and AMS symptoms compared to acute exposure. Finally, a second identical high-altitude sojourn after a week near sea level will be associated with higher SpO_2_ and lower AMS severity during acute and prolonged exposure compared to the first altitude ascent.

## 2. Methods

### 2.1. Study design and setting

This prospective cohort study was conducted in Santiago de Chile (520 m), the ALMA Operation Support Facility (OSF, 2900 m), and the ALMA Operation Site (AOS, 5050 m). Previous results of this project were published elsewhere, however, findings shown in this article have not been previously published (Furian et al., 2022; Graf et al., 2022; Lichtblau et al., 2021; Pun et al., 2019; Pun, Guadagni, et al., 2018; Pun, Hartmann, et al., 2018, 2018).

The study comprises two multi-day cycles during which participants spent 7 days at high and very high altitude (2900 m, respectively 5050 m) interrupted by a recovery period of 7 days in Santiago de Chile (520 m). The participants spent the first days in Santiago de Chile, where baseline measurements were performed. Thereafter, they travelled by commercial airline to San Pedro de Atacama (2 hours) followed by ground transportation (1 hour) to 2900 m for the first altitude sojourn cycle. The participants spent the nights at 2900 m and travelled each day by car (45 min) to 5050 m, where they stayed for 4-8 h without oxygen supplementation or medication. The same measurements as during the baseline at 520 m were carried out at 2900 m in the morning and evening, and at 5050 m at midday. After the 7-day-long recovery phase, the second cycle was conducted with the same altitude exposure pattern as in the first cycle (**Figure 1**).

**Figure 1.**
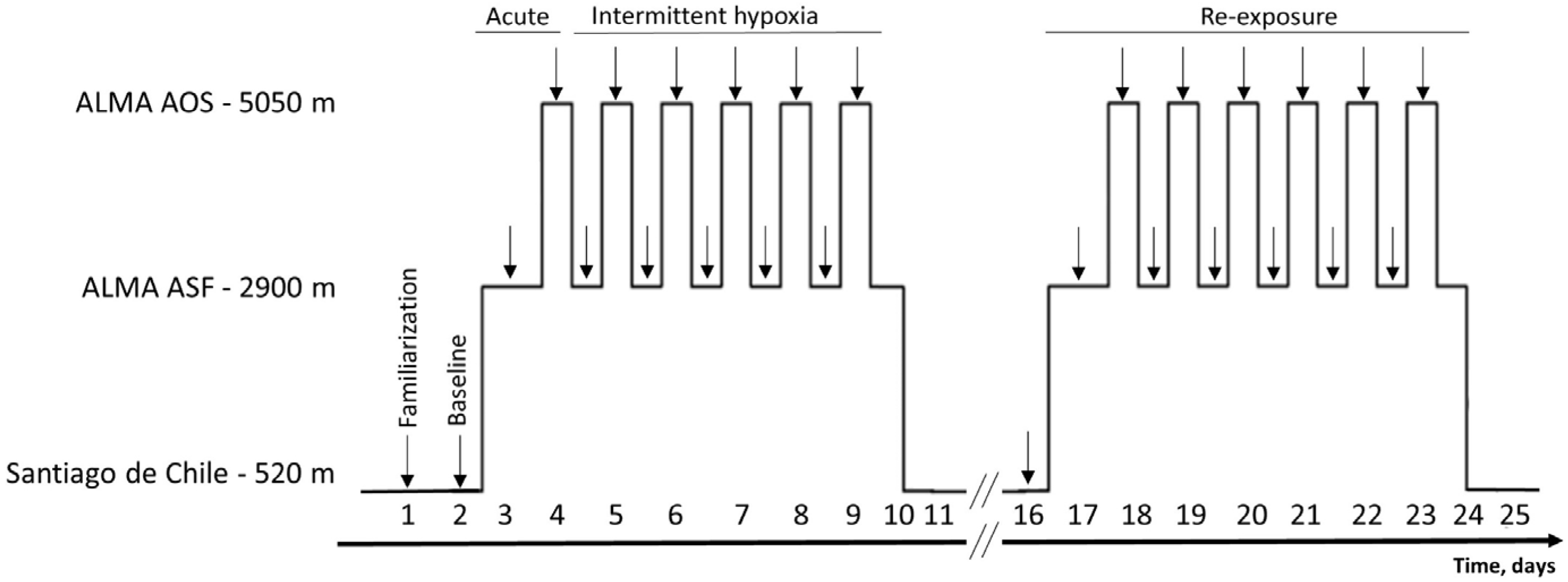
Study setting. In the first three days the participants underwent familiarization and baseline measurement at Santiago de Chile (520 m) before they travelled to ALMA ASF (2900 m) for the first sojourn (Cycle 1). They spent 6 nights at 2900 m where they underwent measurements. During the day, the participants travelled to ALMA AOS (5050 m) where the same measurements were performed. This 7-day cycle at altitude was repeated after a 7-day recovery phase in Santiago de Chile. Arrows indicate the timing of assessments.

### 2.2. Participants

Study participants, aged between 18 to 30 years, were recruited in Calgary (Canada) and Zurich (Switzerland). Exclusion criteria were a positive history of intolerance to altitude (<3000 m), pregnancy, and acute or chronic health conditions requiring medical treatment. Overnight stays at altitudes >1500 m must have been avoided within four weeks prior to the initial study. During the study, alcohol consumption and medication intake were prohibited. The study was approved by the Conjoint Health Research Ethics Board of the University of Calgary (Ethics ID: REB 15-2709) and the Cantonal Ethics Committee of Zurich (2016-00048). The main trial was registered at ClinicalTrials.gov (NCT02731456).

### 2.3. Protocol and measurement

The study started with a familiarization session in Santiago de Chile, followed by baseline measurements. The participants underwent clinical examination, including a morning measurement of body weight and height and three daily measurements of resting SpO2, resting blood pressure, and resting heart rate (morning, midday, and evening). Brachial blood pressure and heart rate were always measured in triplicates while sitting, and the mean of the last two measurements was calculated. Additionally, the participants completed questionnaires to determine the severity of AMS symptoms using the Environmental Symptom Questionnaire – cerebral score (AMSc) in the morning and at midday. The AMSc contains items related to AMS symptoms such as lightheadedness, headache, dizziness, feeling faint, dim vision, coordination disturbances, weakness, nausea, loss of appetite, sickness, and feeling hung over. Each of these symptoms is scored from 0 (not at all) to 5 (very severe). The AMSc is calculated by multiplying every symptom by its factorial weight and summing them up. Per definition, the participants experienced AMS when the AMSc reached scores ≥0.7 (Sampson et al., 1983).

### 2.4. Outcomes

The main outcomes were SpO_2_ and AMSc during acute (defined as the first 12 hours at the target altitude), prolonged intermittent (defined as a 7-day exposure), and repeated (after a 1-week washout period at sealevel) altitude exposure to 2900 and 5050 m, respectively. Secondary outcomes were heart rate and systemic blood pressure.

### 2.5. Statistical analyses

This is a post-hoc analysis of secondary outcomes. After visual inspection of the data distribution using Kernel density plots, mean, standard deviation, and mean differences, including 95% CI, were computed using multi-level mixed-effects linear regression analyses. Main outcomes were assigned as dependent variables (SpO_2_, AMSc), and altitude (520 m, 2900 m, 5050 m), altitude sojourn (Cycle 1, Cycle 2), and day at altitude (1 to 7) as well as their interaction as fixed effects. Participants were included as random effects in these models. Missing data were not replaced. Statistical significance was defined as P <0.05 or the 95%CI, not including the value zero. Statistical analyses were performed with STATA version 15. Unless otherwise stated, values are reported as mean ± SD.

## 3. Results

### 3.1. Study subjects

All recruited participants (n=21) were included in the final statistical analyses. There were no adverse events excluding participants or study withdrawals. The demographic characteristics are presented in **Table 1** . Overall, 13 (61.9%) of the participants were females with a mean ± SD age of 24.6 ± 3.7 years and a body mass index of 22.4 ± 3.0 kg/m .

**Table 1.** Participant characteristics.

|  |  |
| --- | --- |
| N | 21 |
| Age [years] | 24.6 ± 3.7 |
| Weight [kg] | 64.7 ± 9.0 |
| Height [m] | 1.7 ± 0.1 |
| Body mass index [kg/m<sup>2</sup>] | 22.4 ± 3.0 |

Values are presented as counts respectively means ± standard deviation.

### 3.2. Effect of acute, intermittent, and repeated altitude exposure on SpO_2_ and AMS

The effect of acute altitude exposure was measured on the first day at 2900 and 5050 m, respectively, and compared to 520 m. The results are illustrated in **Figure 2** and outlined in **Table 2**. Baseline measurements at 520 m revealed a mean SpO_2_ of 97.6 ± 3.2% and an AMSc score of 0.05 ± 0.6. With ascent to 2900 m, SpO_2_ decreased to 93.7 ± 4.4% (P<0.05 vs. 520 m) and AMSc score increased to 0.44 ± 0.86 (P<0.05 vs. 520 m). The effects of decreasing SpO_2_ and increasing AMS symptoms were more pronounced with acute exposure to 5050 m (decrease of SpO_2_ to 80.0 ± 4.4%; increase of AMSc to 0.98 ± 0.9) (P<0.05 vs. 520 m and vs. 2900m, **Table 2**). Overall, 5 (23.8%) and 12 (57.1%) subjects developed AMS at 2900 and 5050 m (P<0.05, Fisher Exact Test), respectively.

**Figure 2.**
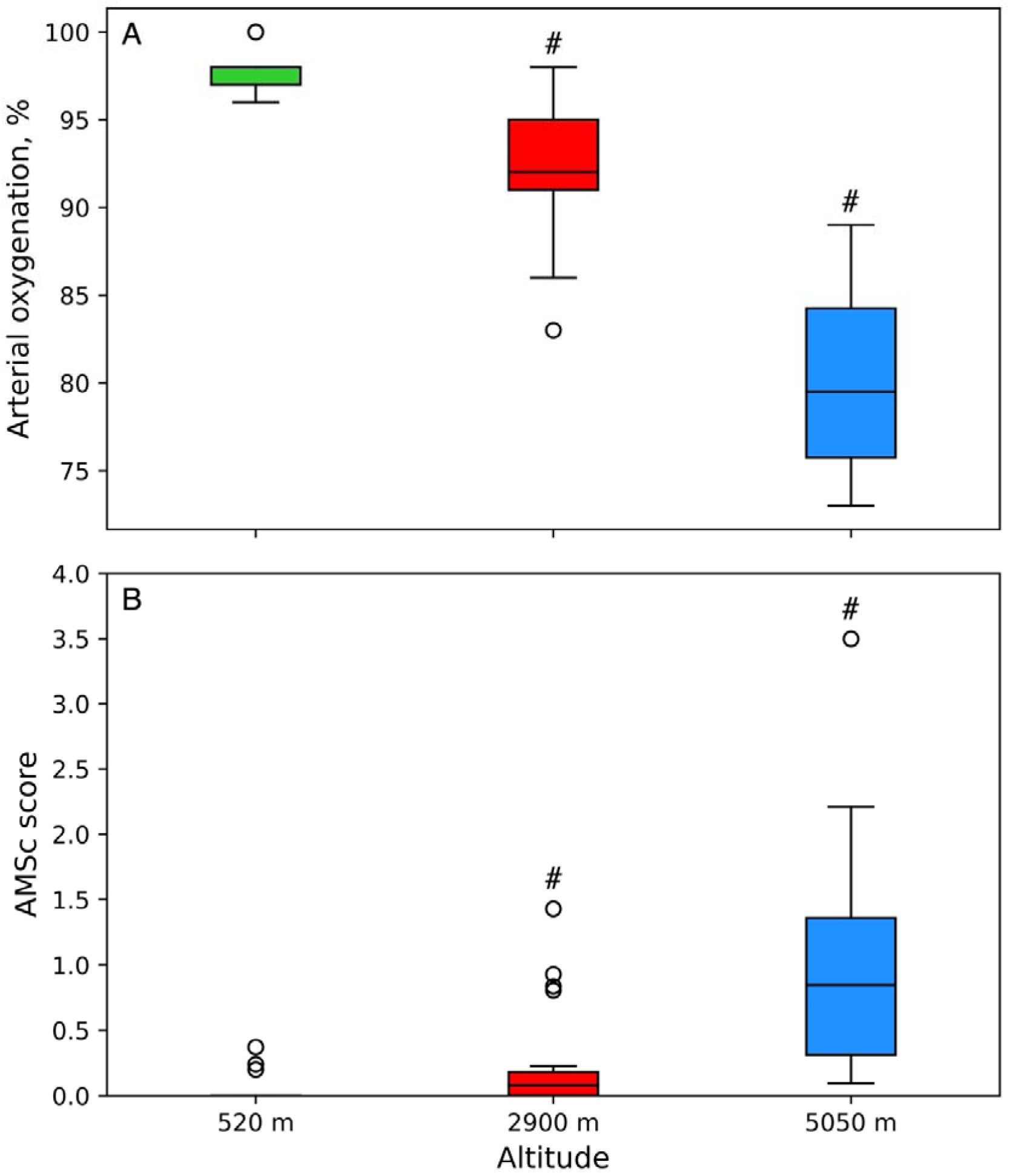
Acute altitude exposure on arterial oxygen saturation and AMS symptoms. Effect of acute altitude exposure on arterial oxygen saturation (A) and AMS symptoms (B). Values are presented as medians and interquartile ranges, the whiskers indicate the 95%-CI. P <0.05 compared to 520 m.

**Table 2.** Acute altitude exposure.

| Altitude | 1<sup>st</sup> exposure to high altitude (Cycle 1) |  |  | 2<sup>nd</sup> exposure to high altitude (Cycle 2) |  |  |
| --- | --- | --- | --- | --- | --- | --- |
|  | 520 m | 2900 m | 5050 m | 520 m | 2900 m | 5050 m |
| SpO<sub>2</sub>, % | 97.6 ± 3.2 | 93.7 ± 4.4* | 80.0 ± 4.4*<sup>#</sup> | 98.1 ± 2.5 | 94.1 ± 4.3* | 82.8 ± 4.3*<sup>#†</sup> |
| AMSc | 0.05 ± 0.63 | 0.44 ± 0.86* | 0.98 ± 0.87*<sup>#</sup> | 0.06 ± 0.22 | 0.06 ± 0.38<sup>†</sup> | 0.60 ± 0.38*<sup>#†</sup> |
| Heart rate, bpm | 65 ± 15 | 81 ± 20* | 98 ± 23*<sup>#</sup> | 57 ± 10<sup>†</sup> | 74 ± 15*<sup>#</sup> | 95 ± 15*<sup>#</sup> |
| Systolic BP, mmHg | 114 ± 13 | 114 ± 18 | 120 ± 19<sup>#</sup> | 104 ± 12<sup>†</sup> | 112 ± 17* | 120 ± 17*<sup>#</sup> |
| Diastolic BP, mmHg | 69 ± 9 | 71 ± 12 | 77 ± 13*<sup>#</sup> | 61 ± 10<sup>†</sup> | 70 ± 14* | 77 ± 14*<sup>#</sup> |

Values are presented in means ± SD. \*P <0.05 compared to 520 m; <sup>#</sup>P<0.05 compared to 2900 m; <sup>†</sup>P <0.05 compared to the value of the corresponding altitude in Cycle 1. SpO<sub>2</sub>, arterial oxygen saturation assessed by finger pulse oximetry; AMSc, acute mountain sickness score assessed by the Environmental Symptom Questionnaire – cerebral score. An AMSc score of ≥0.7 represents clinically relevant acute mountain sickness; BP, blood pressure.

The effects of prolonged intermittent altitude exposure to 2900 m and 5050 m on SpO_2_ and AMS symptoms are displayed in **Figure 3** and **Table 2**. In Cycle 1, SpO_2_ increased at 2900 m by a mean (95% CI) of 0.3 %/day (0.2 to 0.4) and at 5050 m by 0.9 %/day (0.4 to 1.3); while the AMSc decreased at 2900 m by 0.05 points/day (0.01 to 0.08) and at 5050 m by 0.16 points/day (0.11 to 0.21).

**Figure 3.**
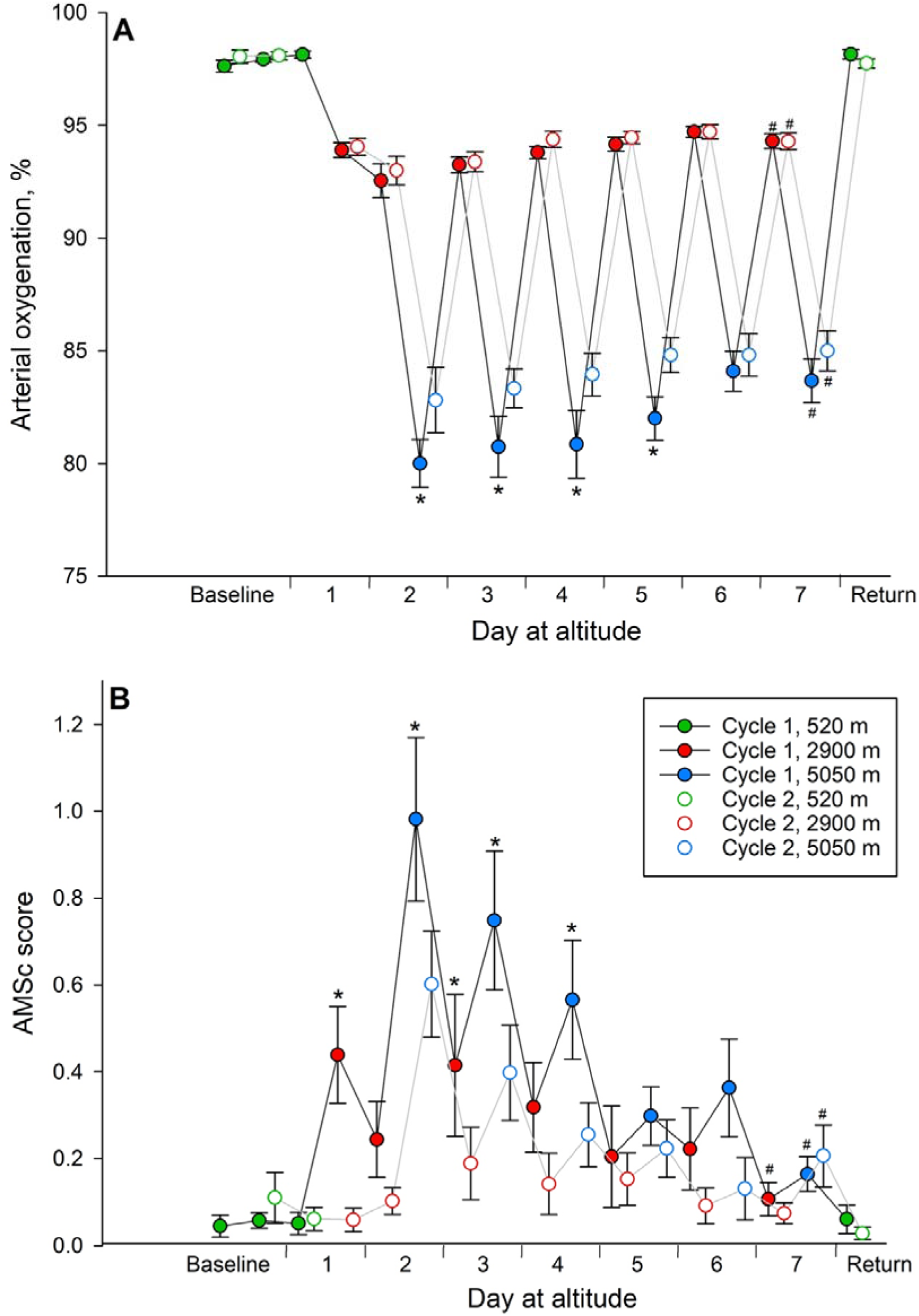
Effect of intermittent hypoxia and repeated altitude exposure on arterial oxygen saturation and AMS symptoms. Effect of intermittent hypoxia and altitude re-exposure on arterial oxygen saturation (A) and AMS symptoms (B). Baseline values were measured at 520 m. The dots represent mean values and the error bars the standard deviation. The number sign (#) represents p<0.05 on the following days compared to the first day at 2900 m respectively 5050 m (prolonged exposure); the asterisk (*) represents p<0.05 on the corresponding days at 2900 m respectively 5050 m in Cycle 2 vs. Cycle 1 (re-exposure).

The effects of repeated altitude exposure on SpO_2_ and AMS symptoms were analyzed by comparing the same day at altitude of Cycle 2 vs. that in Cycle 1 at 2900 m and 5050 m, respectively (**Figure 3**). Compared to Cycle 1, the absolute values of SpO_2_ were unchanged at any time point at 2900 m but improved at 5050 m (**Figure 3**, Panel A) during Cycle 2. The AMSc decreased during several days in Cycle 2 at 2900 m and 5050 m compared to the corresponding days in Cycle 1 (**Figure 3**, Panel B). Corresponding changes with intermittent hypoxia during Cycle 2 in SpO_2_ at 2900 m were: 0.3 %/day (0.1 to 0.4) and at 5050 m: 0.5 %/day (0.11 to 0.80); in AMSc at 2900 m: 0.00 points/day (−0.02 to 0.02) and at 5050 m: -0.08 points/day (−0.04 to - 0.12).

In Cycle 2, nobody developed AMS on acute exposure to 2900 m (P<0.05 vs Cycle 1) but 6 (28.6%) participants at 5050 m (P = 0.058 vs Cycle 2).

### 3.3. Effect of acute, intermittent, and repeated altitude exposure on heart rate, systolic and diastolic blood pressure

Results are displayed in **Figure 4** and **Table 2**. Baseline values measured at 520 m revealed a mean heart rate of 65 ± 15 bpm, systolic blood pressure of 114 ± 13 mmHg, and diastolic blood pressure of 69 ± 9 mmHg. At first exposure to 2900 m, there was an increase in heart rate to 81 ± 20 bpm (P<0.05 vs. 520 m) but no change in systolic or diastolic blood pressure. The increase in heart rate was even more pronounced at acute exposure to 5050 m (increase of heart rate to 98 ± 23 bpm) (P<0.05 vs. 520 m and 2900 m, **Table 2**). At 5050 m, systolic blood pressure was higher compared to 2900 m, whereas diastolic blood pressure increased to 77 ± 13 mmHg and was higher compared to 520 m and 2900 m.

**Figure 4.**
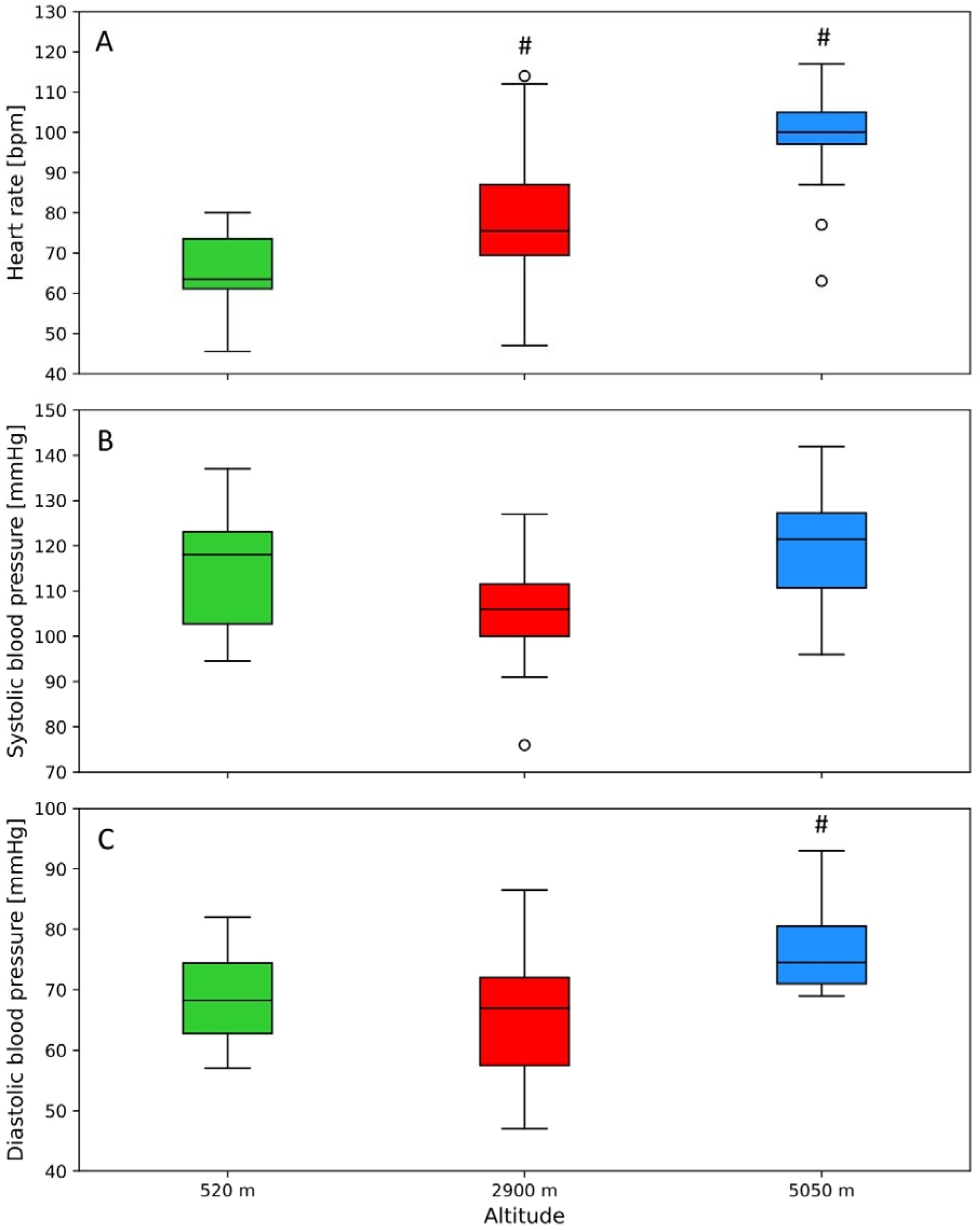
Acute altitude exposure on heart rate, systolic and diastolic blood pressure. Effect of acute altitude exposure on heart rate (A), systolic blood pressure (B) and diastolic blood pressure (C). Values are presented as medians and interquartile ranges, the whiskers indicate the 95%-CI. The number sign (#) indicates p<0.05 on altitude 2900 respectively 5050 m compared to 520 m.

The effects of intermittent altitude exposure on systolic and diastolic blood pressure and heart rate were analyzed by comparing consecutive days at altitude to the 1^st^ day at 2900 m, respectively 5050 m. Results are displayed in **Figure 5**. In Cycle 1, systolic blood pressure decreased at 5050 m by a mean (95% CI) of 2 mmHg/day (0 to 3 mmHg/day) (P<0.05 compared to no change), but not at 2900 m. In Cycle 2, there was only a decrease in systolic blood pressure at 2900 m by 1 mmHg/day (0 to 1 mmHg/day) (P<0.05 compared to no change), but not at 5050 m. The diastolic blood pressure did not change at 2900 m or 5050 m. In Cycle 1, heart rate decreased at 5050 m by 5 bpm/day (2 to 7 bpm/day) as well as at 2900 m by 2 bpm/day (0 to 4 bpm/day) (P<0.05 both comparisons compared to no change). In Cycle 2 there was also a decrease in heart rate at 5050 m by 2 bpm/day (1 to 3 bpm/day) (P<0.05 compared to no change), but not at 2900 m.

**Figure 5.**
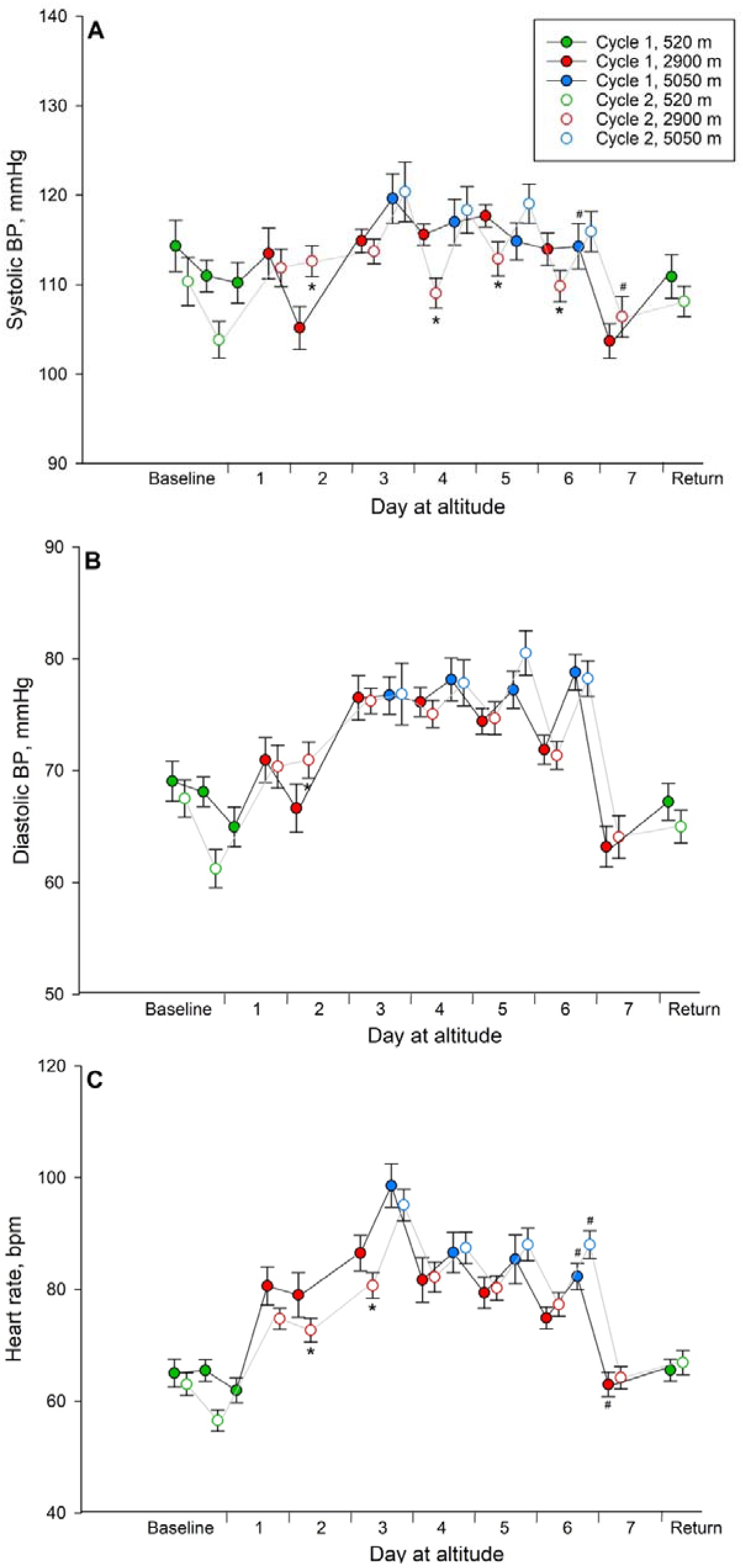
Effect of intermittent hypoxia and repeated altitude exposure on blood pressure and heart rate. Effect of intermittent hypoxia and repeated altitude exposure on systolic blood pressure (A), diastolic blood pressure (B) and heart rate (C). Baseline values were measured at 520 m. The dots represent mean values and the error bars the standard deviation. The number sign (#) represents p<0.05 on the following days compared to the first day at 2900 m respectively 5050 m (prolonged exposure); the asterisk (*)represents p<0.05 on the corresponding days at 2900 m respectively 5050 m in Cycle 2 vs. Cycle 1 (re-exposure).

The effect of a second intermittent-hypoxic exposure on systolic and diastolic blood pressure and heart rate were analyzed by comparing the day of Cycle 2 to the same day of Cycle 1 at 2900 m and 5050 m, respectively. Results are displayed in **Figure 5**. Compared to Cycle 1, absolute systolic blood pressure values were increased in Cycle 2 on day 2 and decreased on days 4 to 6 at 2900 m. Systolic blood pressure remained unchanged at 5050 m (**Figure 5**, Panel A). Diastolic blood pressure was only increased in Cycle 2 on day 2 at 2900 m compared to the corresponding day of Cycle 1. At 5050 m, there was no decrease in diastolic blood pressure present on any day (**Figure 5**, Panel B). Heart rate decreased on day 2 and day 3 in Cycle 2 compared to the corresponding day of Cycle 1 at 2900 m. A decrease in heart rate could not be shown on any measurement at 5050 m (**Figure 5**, Panel C). Corresponding changes during the intermittent hypoxic stay during Cycle 2 in systolic blood pressure at 2900 m were: -1 mmHg/day (−1 to 0 mmHg/day) and at 5050 m: -1 mmHg/day (−3 to 1 mmHg/day); in diastolic blood pressure at 2900 m: -1 mmHg/day (−1 to 0 mmHg/day) and at 5050 m: 1 mmHg/day (−1 to 2 mmHg/day); in heart rate at 2900 m: 0 bpm/day (−1 to 0 bpm/day) and at 5050 m: -1 bpm/day (−4 to 0 bpm/day).

## 4. Discussion

This study in healthy lowlanders ascending to and sleeping at 2900 m and travelling daily to 5050 m for 7 days revealed that intermittent hypoxic exposure increased SpO_2_ and decreased AMS symptoms at both altitudes but faster at 5050 m. In turn, intermittent hypoxia was not associated with changes in heart rate and systemic blood pressure at either altitude. Moreover, a repeated identical intermittent hypoxic exposure pattern after one week of low altitude recovery showed faster high-altitude acclimatization, suggested by less hypoxemia and less AMS symptoms, especially at 5050 m. Although participants showed better altitude tolerance and further improvements in SpO_2_ and AMS symptoms during the second sojourn to high altitudes, no changes in heart rate and systemic blood pressure were observed. These findings suggest beneficial effects of intermittent hypoxic exposure and repeated altitude exposure pattern on oxygenation and AMS symptoms without detectable stress relief in cardiovascular metrics.

### 4.1. Effects of acute altitude exposure

Our findings are consistent with the current knowledge of acute high-altitude exposure. In agreement with previous studies (Cobb et al., 2021; Subudhi, Bourdillon, et al., 2014; West, 2012), SpO_2_ decreased after ascent with an inter-individual variation and with a higher degree at 5050 compared to 2900 m. Correspondingly, AMS symptoms developed mainly during acute exposure to 5050 m (57.1% AMS incidence) but less at 2900 m (23.8% AMS incidence) (**Figure 2**). Hypoxemia increased heart rate and diastolic (not systolic) blood pressure, especially at 5050 m compared to 2900 m (**Figure 4**). Regarding changes in systemic blood pressure during exposure to high altitude, the literature shows inconsistent findings. While several studies reported a continuous increase in systemic blood pressure proportional to the altitude reached (Bärtsch & Saltin, 2008; Bilo et al., 2019; Cobb et al., 2021; Paralikar & Paralikar, 2010; Vogel et al., 1974; Wait et al., 2023), a few studies could not find a blood pressure increase with altitude. This is explainable by systemic hypoxic vasodilation shortly after acute exposure and consequently transient decrease in blood pressure within the first hours of exposure (Naeije, 2010). The lack of blood pressure increase in our study might be related to sample size or to study design differences (i.e., different absolute altitudes reached, blood pressure assessment techniques (spot checks vs. 24-hour ambulatory blood pressure measurement), daytime variations or differences in postural position (supine, sitting and standing) or other, unknown factors).

### 4.2. Effects of prolonged intermittent altitude exposure

The SpO_2_ increased during the 7-day prolonged intermittent stay at 2900 m and 5050 m (**Figure 3**, Panel A). Similar observations were noted after an ascent from sea level to 4300 m (Reeves et al., 1993). With prolonged intermittent altitude exposure, SpO_2_ increased, but baseline values could not be reached. This finding correlates with that of previous studies (Subudhi, Bourdillon, et al., 2014; Vogel et al., 1974). Furian et al. investigated in the same study population nocturnal breathing and found that nocturnal SpO_2_ rose during the stay at high altitude, what suits our findings of diurnal increase in SpO_2_ (Furian et al., 2022). In our study, SpO_2_ rose by 0.3% per day of acclimatization at 2900 m and 0.9% per day at 5050 m, while at night, SpO_2_ rose by approximately 0.7%/day at 2900 m. This observation of nocturnal SpO_2_ increment is within the observed daytime SpO_2_ increments at 2900 m and 5050 m of this study and confirms that SpO_2_ increments correlate with the severity of nocturnal hypoxemia. In another study conducted by Subudhi et al., they found in 21 lowlanders staying at 5260 m for 16 days, an approximate arterial oxygen saturation (SaO_2_) increase of 0.3%/day (calculated from the presented mean SpO_2_ values of the article) (Subudhi, Bourdillon, et al., 2014). We observed a greater increase of SpO_2_ at 5050 m compared to 2900 m with acclimatization, which might be explained by a larger hypoxic stress and proportionally higher hypoxic ventilatory response. The ventilatory response is regulated by oxygen sensing chemoreceptors in the carotid bodies (Vassilakopoulos, 2012). Only minimal hypoxic ventilatory response occurs above SpO2 values of >90%. Below this threshold, a sharp increase in hypoxia-related hyperventilation occurs (Vassilakopoulos, 2012). In accordance, SpO_2_ was always >90% at 2900 m. Thus, hypoxic ventilatory response might have been minimal, whereas, at 5050 m, SpO2 was 80.0 ± 4.4%, presumably leading to increased ventilation and consequently improving with intermittent hypoxic exposure.

In our study, AMSc decreased throughout the stay at high altitude (**Figure 3**, Panel B). This was also found by Imray and colleagues (Imray et al., 2010), who described similar findings with fewer occurrences of AMS after the initial hypoxic exposure and with the onset of acclimatization. Additionally, Subudhi et al. (Subudhi, Bourdillon, et al., 2014) found that AMS symptoms decreased during a 16-day stay at 5260 m and were completely resolved at the end of the sojourn.

Heart rate decreased with prolonged intermittent hypoxia in the first Cycle at 5050 m and 2900 m but stayed elevated compared to baseline values (**Figure 5**, Panel C). These observations align with those of prior investigations (Milledge et al., 1983; Vogel et al., 1974) and heart rate values obtained at night in the same study population (Furian et al., 2022). Similarly, systolic blood pressure decreased at 5050 m over time while diastolic blood pressure remained elevated (**Figure 5**, Panel A and B). These findings are somewhat in contrast to the previously reported persistent elevation of systemic blood pressure at 5400 m (Bilo et al., 2019). Whether the decrease in systolic blood pressure can be assigned to the intermittent exposure to 2900 and 5050 m remains to be elucidated.

### 4.3. Effects of re-exposure to high altitude

With acute re-exposure to high altitude, SpO_2_ values were higher at 5050 m but unchanged at 2900 m compared to acute exposure in Cycle 1 (**Figure 3**, Panel A). Similarly, an improved SpO_2_ could be seen in subjects with previous altitude exposure climbing from Lukla (2840 m) to Gokyo Ri (5360 m) in Nepal compared to altitude-naïve subjects (MacNutt et al., 2012). Further studies, which have reported that intermittent altitude exposure resulted in elevated SpO_2_ values during re-exposure to high altitude, support our findings. For example, Subudhi et al. reported improved mean SaO_2_ at 5260 m from 76.4% at first exposure to 81.6% and 82.4% at re-exposure after 7 and 21 days recovery at 1500 m, mean SaO_2_ was 81.6% respectively 82.4% compared to 76.4% at first exposure (Subudhi, Bourdillon, et al., 2014). Beidleman et al. demonstrated that intermittent altitude exposure to 4300 m improved SaO_2_ from an initial 80% at first acute exposure to 4300 m to 85% after intermittent altitude exposure thereafter (Beidleman et al., 2004). These findings support ours, which revealed a SpO_2_ of 80.0% at first exposure to 5050 m and 82.8% at re-exposure. Although our investigations could not show a change in diurnal mean SpO_2_ during re-exposure to 2900 m (93.7% at first and 94.1% at re-exposure), nocturnal changes of SpO_2_ at 2900 m with re-exposure, which were reported previously, were slightly improved (86% to87%, P<0.05). (Furian et al., 2022). Thus, our findings of increasing SpO_2_ with re-exposure to 5050 m but not to 2900 m align with previous investigations.

In accordance with improved SpO_2_ values, re-exposure led to significantly lower AMSc during the second compared to the first sojourn (**Figure 3**, Panel B). These lower AMScs were probably caused by better oxygenation, a carry-over, or a memory effect. These observations were in accordance with previous studies showing that frequent altitude exposure and greater previous maximum target altitude were associated with a lower prevalence of AMS (Beidleman et al., 2004; Imray et al., 2010; MacNutt et al., 2012; Pesce et al., 2005; Schneider et al., 2002; Weng et al., 2013). However, even with prolonged intermittent altitude exposure and re-exposure after only 1 week near sea-level, AMS could not fully be prevented. These findings correlate with those of a study investigating workers of the Collahuasi mine in Chile sleeping at 3800 m and working at 4600 m with a similar working schedule as those of the ALMA observatory. Richalet et al. found in these miners no significant decrease in AMS with persistent exposure but better oxygen saturation (J. P. Richalet et al., 2002). Although, in the hypobaric chamber, AMS symptoms were reduced during reexposure and could almost be prevented up to a pressure of 5000 m thanks to pre-acclimatization (J.-P. Richalet et al., 1999). Furthermore, Subudhi et al. (Subudhi, Bourdillon, et al., 2014) studied retention of acclimatization after 16 days at 5260 m and found that subjects were still protected from AMS after a 7-day recovery period at 1525 m whereas the effect was not sustained after 21 days at lower altitude.

Within secondary outcomes, heart rate decreased at 2900 m but remained unchanged at 5050 m during acute exposure in Cycle 2 compared to Cycle 1 (**Figure 5**, Panel C). These findings require further investigation and cannot be explained by the improved SpO_2_ at 2900 and 5050 m. Notably, other studies who looked at the changes of heart rate during re-exposure to high altitude were also not able to show a significant decrease (Beidleman et al., 2004; MacNutt et al., 2012).

In our study, re-exposure to high altitude resulted in an inconsistent change in systolic and diastolic blood pressure (**Figure 5**, Panel A and B). A comparable study examining Chilean miners with a similar altitude exposure profile showed a tendency that systemic blood pressure decreased with time in subjects with chronic intermittent altitude exposure but remained above baseline values (J. P. Richalet et al., 2004). However, during other investigations, re-exposure to high altitude did not change blood pressure (Beidleman et al., 2004, 2009; MacNutt et al., 2012).

### 4.4. Strengths and limitations

The strength of this project was the repeated assessments at 2900 m and 5050 m, which corresponds to the pattern of real-life high-altitude workers and the relatively homogenous population. Limitations of the current report might be the post-hoc analysis of the data, the small sample size not allowing the assessment of sex differences in the acclimatization and repeated altitude exposure effects, and the lack of the Lake Louise Questionnaire scores for AMS.

## 5. Conclusion

In this prospective cohort study, we present comprehensive insights on the effect of prolonged intermittent exposure to high and very high altitudes and the effects of re-exposure to high altitudes on SpO_2_, AMS, and cardiovascular parameters. Such a “sleep-high work-very-high” working schedule is representative of many high-altitude workers worldwide. We found that SpO_2_ and AMS symptoms improved over the 7-day intermittent high-altitude exposure, especially at 5050 m, less at 2900 m. Full acclimatization never occurred, however, a second sojourn, and identical intermittent high altitude pattern resulted in faster altitude tolerance at 5050 m. These findings are important to consider when newly recruited high-altitude workers commence work. Nevertheless, despite improvements in SpO_2_ and AMS symptoms, no improvements in cardiovascular metrics were observed. Further studies on long-term high-altitude workers are warranted.

## Data Availability

All data produced in the present study are available upon reasonable request to the authors.

